# Rationale and design of the PREGnancy, HEART Health and Cardiovascular Disease (PREG-HEART) Cohort Study

**DOI:** 10.64898/2026.03.26.26349373

**Authors:** Kathryn Hunt, Rachel Buchan, UK Maternal Cardiovascular Health Collaborative Group, Claire Sheppard, Rachel Cartwright, Simon Fisher, Rebecca Jarman, Rebecca M Reynolds, James S Ware, Tim Chico, Deborah A Lawlor, Antonio de Marvao, Upasana Tayal

**Affiliations:** Centre for Cardiovascular Science, University of Edinburgh, Edinburgh, UK; National Heart and Lung Institute, Imperial College London, London, UK; Medical Research Council Laboratory of Medical Sciences, Imperial College London, London, UK; Royal Brompton and Harefield Hospitals, Guy’s and St Thomas’ NHS Foundation Trust, London, UK; NIHR-BHF Cardiovascular Partnership, University of Oxford, Oxford, UK; Guy’s and St Thomas’ NHS Foundation Trust; British Heart Foundation Data Science Centre, Health Data Research UK, London, UK; NIHR Sheffield Biomedical Research Centre, Sheffield Teaching Hospitals NHS Foundation Trust and University of Sheffield, Sheffield, UK; Medical Research Council Integrative Epidemiology Unit, University of Bristol, Bristol, UK; Population Health Science, Bristol Medical School, University of Bristol, Bristol, UK; Department of Women and Children’s Health, School of Life Course & Population Sciences, King’s College London, UK; British Heart Foundation Centre of Research Excellence, School of Cardiovascular and Metabolic Medicine & Sciences, King’s College London, London, UK

## Abstract

**Introduction:** Cardiovascular disease is a leading cause of maternal and neonatal morbidity and mortality in the UK. Its prevalence in pregnancy continues to rise, driven by both improved survival of women with congenital and inherited heart disease into reproductive age and an increasing burden of acquired cardiovascular risk factors. However, its natural history and optimal management remain poorly defined. Current research is limited by small sample sizes, drawn from highly selected patient cohorts from individual units. The aim of the PREGnancy, HEART Health, and Cardiovascular Disease (PREG-HEART) study is to develop a patient driven, clinically relevant, digital platform to understand the epidemiology of cardiovascular disease in pregnancy and support clinical trials of management strategies. This paper provides the protocol for PREG-HEART, which will start with a 6-month pilot study.

**Methods and analysis:** PREG-HEART will utilise an online, direct-to-patient platform to enrol patients with cardiovascular disease in pregnancy alongside healthy pregnant controls. Enrolled women will be invited to provide self-reported demographic and clinical data and consent to linkage with national health records for long-term follow up. We will also seek consent for storage and analysis of leftover clinical biosamples and to re-contact participants, enabling recruitment into sub-studies and clinical trials.

Planned analysis for the pilot study at 6 months will assess feasibility, including recruitment rates, case-mix of cardiovascular diagnoses, and participant geographical, socio-economic, and ethnic background compared to the UK general pregnant population. Findings from the pilot study will inform subsequent phases of PREG-HEART, which will explore associations between different cardiovascular diagnoses and adverse cardiovascular, obstetric, and neonatal events. We will work closely with patients and clinicians to define priority research questions and use the PREG-HEART platform to support a range of observational and interventional studies to address these.

**Ethics:** This study was approved by the West Midlands Solihull Research Ethics Committee.

**Registration details:** PREG-HEART has been registered prospectively on the ISRCTN registry (ISRCTN11700499)

**WHAT IS ALREADY KNOWN ON THIS TOPIC:**

- Cardiovascular disease in pregnancy is a leading cause of adverse health outcomes for both mothers and babies in the UK and its prevalence is rising.
- Data to guide management of cardiovascular disease in pregnancy are limited, including how to care for women during pregnancy and the postpartum, the impact of maternal disease on pregnancy outcomes, and the effect of pregnancy on long-term cardiovascular health.
- Because individual conditions (e.g. specific cardiomyopathies or congenital lesions) are rarely encountered within single maternity units, existing studies are small, underpowered, and restricted to highly selected patient groups from a small number of large units.

**WHAT THIS STUDY ADDS:**

- PREGnancy, HEART Health, and Cardiovascular Disease (PREG-HEART) is a planned national, prospective, data-enabled cohort and platform study.
- Women with cardiovascular disease in pregnancy and healthy pregnant controls will be recruited via a direct-to-patient online platform, allowing entry of self-reported demographic and clinical data, consent to long-term data linkage, consent to recontact for additional studies and biosample collection.
- This direct-to-patient approach is designed to overcome geographical and institutional barriers, ensuring that the cohort is diverse and representative of the wider UK population.
- In an initial pilot phase, we will assess the feasibility of recruitment to PREG-HEART and describe the representativeness of our cohort with regards to the geographical, socio-economic and ethnic background of participants compared with the UK pregnant population.
- The results of this pilot will inform a larger study which will leverage additional funding and utilise the PREG-HEART platform to enable aetiological studies and clinical trials that answer priority questions of patients and health care providers.

**HOW THIS STUDY MIGHT AFFECT RESEARCH, PRACTICE, OR POLICY:**

- We anticipate that this pilot study will lay vital groundwork and demonstrate the feasibility of direct-to-patient recruitment to an online pregnancy cohort.
- This will inform future work to develop PREG-HEART as a unique national resource to support collaborative clinical and translational research, strengthen epidemiological evidence, and inform future strategies for the management of cardiovascular disease in pregnancy.

**Graphical Abstract:** 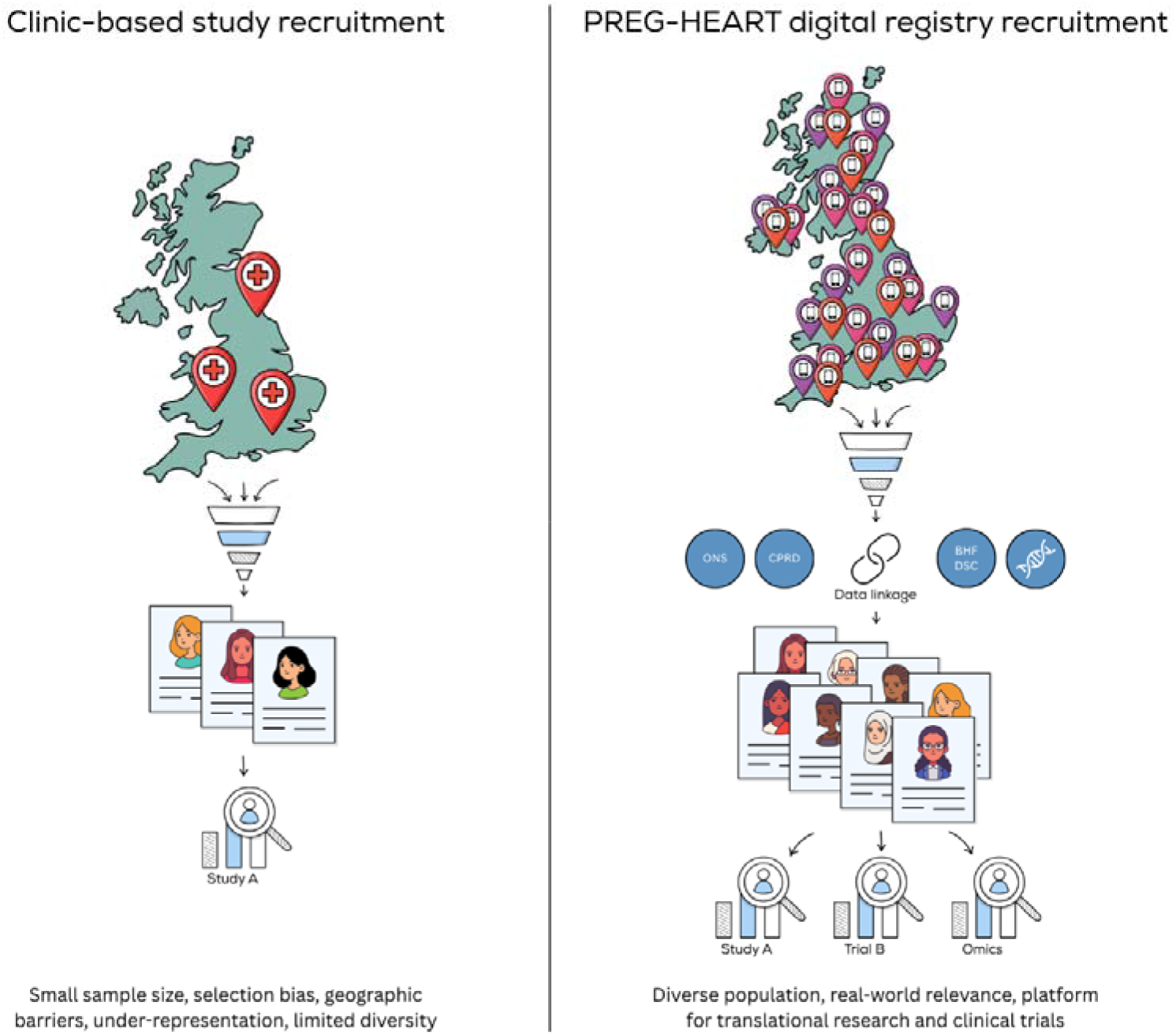

## Introduction

Cardiovascular disease is a leading cause of maternal morbidity and mortality in the UK, and a major contributor to adverse fetal and neonatal outcomes(1–3). Up to 4% of pregnant women have cardiovascular disease and this prevalence is increasing, driven both by improved survival into adulthood of women with inherited and congenital heart diseases, and by the rising burden of acquired risk factors for cardiovascular disease, such as advanced maternal age, obesity, diabetes mellitus, and hypertension(4).

There has been no significant improvement in maternal death rates from cardiovascular disease over recent decades, and outcomes in the UK compare poorly to other high-income countries(1,5). Moreover, there is stark inequality in women’s maternal health outcomes in the UK, with women of Black ethnicity nearly three times more likely than White women to die in childbirth, and Asian women almost twice as likely(1). Maternal mortality is also twice as high in the most socio-economically deprived areas of the UK, compared with the most affluent(1).

Although the cumulative incidence of cardiovascular disease in pregnancy is rising, the relative rarity of individual diagnoses limits research into specific conditions in pregnancy. The natural history of cardiovascular diseases in pregnancy, institutional and regional variation in management, and the relative contributions of genetic and environmental factors remain poorly characterised, largely because sufficiently large cohorts are difficult to assemble. Interventional trials are typically small, underpowered, and restricted to highly selected patient groups(4).

National data collection on cardiovascular disease in pregnancy has up to now relied on clinicians to identify patients and submit data to centralised registries. Such an approach is limited, as it places pressure on busy clinicians, is biased towards better resourced units, and restricts opportunities for participation. For example, the Registry Of Pregnancy And Cardiac disease (ROPAC) study across Europe recruited fewer than 6,000 women over 10 years(6), whereas our preliminary work identified 29,890 women entering pregnancy with previously diagnosed cardiac disease during 2019-2024 in England alone(7). An overreliance on clinician-directed registries also results in marked geographical variation in research access, particularly disadvantaging women who live far from large academic centres.

These factors have direct impacts on clinical care; for example, risk stratification tools which guide patient counselling and management may be based on data which is either outdated or poorly representative of real-world populations. Regional disparities in research participation compound systemic inequities, particularly for groups historically underrepresented and underserved by cardiovascular research(8).

Maternal cardiovascular health is identified as a priority area, and in 2022 was adopted as a flagship theme by the NIHR-British Heart Foundation (BHF) Cardiovascular Partnership. The Partnership is a strategic initiative and NIHR Translational Research Collaboration (TRC) representing 19 member centres (including 11 NIHR Biomedical Research Centres, nine BHF Centres of Research Excellence and four Scottish cardiovascular centres), that facilitates UK-wide cardiovascular research at scale.

Within this framework, the PREGnancy, HEART-Health and Cardiovascular Disease (PREG-HEART) study has been conceived and designed, as a national, direct-to-patient platform. Its overarching aim is to build a large, representative cohort of patients with cardiovascular disease in pregnancy alongside healthy pregnant controls, to enable improved understanding of the contemporaneous natural history of cardiovascular disease in pregnancy, enable robust epidemiological research, and provide a scalable resource for clinical trials and biomarker discovery. By adopting a direct-to-patient approach to recruitment, we seek to overcome the geographical and socio-economic barriers which have driven the underrepresentation of women – particularly pregnant women – in cardiovascular research(9).

### Study Aims

The PREG-HEART study will begin as a pilot phase, which will aim to demonstrate the feasibility of direct-to-patient recruitment for studying cardiovascular disease in pregnancy. At 6 and 12 months, we will assess recruitment rates, case-mix of cardiovascular diagnoses amongst participants, and the sociodemographic, geographical, and ethnic background of participants to determine which, if any, groups are underrepresented by our approach.

Findings from the pilot study will inform further development of the PREG-HEART platform as a patient driven resource with the ability to establish a large, representative, heterogenous, deeply-phenotyped patient cohorts with linkage to obstetric, cardiovascular, and long-term outcomes. This will help build national capacity for cardiovascular research and support observational and interventional studies to address priority research questions for patients and clinicians.

Overall study aims and objectives are summarised in box 1.

#### Box 1: Aims of PREGnancy, HEART Health and Cardiovascular Disease (PREG-HEART) Study

**Overarching aim**

To establish a large, representative, and deeply-phenotyped cohort of women with cardiovascular disease in pregnancy, alongside healthy pregnant controls, to provide a national platform for collaborative clinical and translational research.

**Pilot study objectives**

- Feasibility: Demonstrate that a direct-to-patient online platform can effectively recruit participants with cardiovascular disease in pregnancy.
- Representation: Assess the geographical, socio-economically, and ethnic diversity of recruited participants and identify underrepresented groups.

**Long-term objectives of scaled-up study**

- Natural history and management: Characterise the natural history of different forms of cardiovascular disease in pregnancy and evaluate regional and institutional variation in management practices and outcomes across the UK.
- Bioresource: Create a linked bioresource of clinical samples (blood, saliva, urine) to support multiomic research, improve understanding of pathophysiological mechanisms, and develop biomarkers for diagnosis and risk stratification.
- Platform for trials: Overcome historic, geographical, and institutional barriers to research participation by providing diverse patient groups, with linked outcome data, that are readily available for recruitment into clinical trials.

## Methods and Analysis

### Study design

Study design and participant flow are illustrated in figure 1.

**Figure 1:**
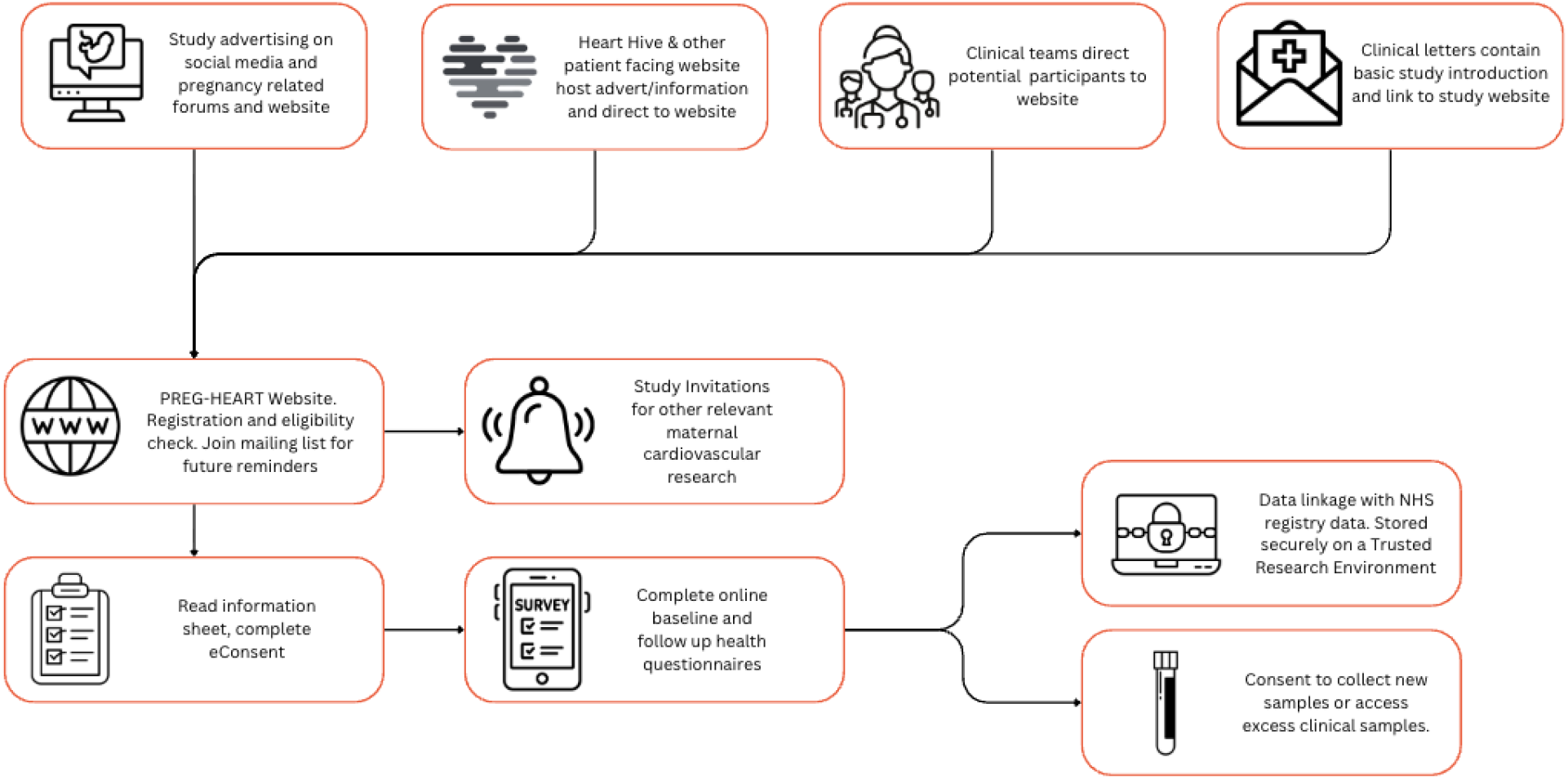
PREGnancy, HEART Health and Cardiovascular Disease (PREG-HEART) study design

PREG-HEART is a bespoke patient-facing, online, observational, prospective study which will enrol participants with cardiovascular disease in pregnancy (cases) and healthy pregnant women (controls) on an ongoing basis. The study was co-designed with a patient steering group and builds upon existing infrastructure of the Heart Hive (https://thehearthive.org), an established patient-facing platform for cardiomyopathy and myocarditis research(10).

### Oversight and governance

The study is sponsored by Imperial College London. Study design and delivery are informed by a multidisciplinary working group from the Maternal Cardiovascular Health Theme of the NIHR-BHF Cardiovascular Partnership, which includes patient representatives and experts in clinical trials, data science, and data governance (membership provided in Appendix 1). An Executive Steering Committee drawn from this working group will be responsible for ongoing governance and oversight of the study.

### Funding and registration

Set-up and initial recruitment to the pilot study is funded by pump-priming funding from the NIHR-British Heart Foundation (BHF) Cardiovascular Partnership. PREG-HEART has been prospectively registered on the ISRCTN clinical study registry (ISRCTN11700499).

### Patients

Eligibility criteria are summarised in box 2. The study is being conducted in accordance with the principles of Good Clinical Practice and the World Medical Association Declaration of Helsinki. All participants will provide electronic informed consent via an online step-by-step consent process on the PREG-HEART web platform.

#### Box 2: Eligibility criteria for the PREGnancy, HEART Health and Cardiovascular Disease (PREG-HEART) Study

**Inclusion criteria**

○ Aged 16 years or above
○ Capacity to provide informed consent

For those enrolling as participants with cardiovascular disease (cases):

○ Any history of pregnancy, either current or previous (with pregnancy defined as a positive pregnancy test and/or ultrasound confirmed pregnancy)
○ Self-reported confirmed diagnosis of cardiovascular disease which predates pregnancy, or was newly diagnosed during pregnancy or within six months postpartum, including:
  ◼ Ventricular dysfunction:
    – cardiomyopathies (inherited or acquired, including peripartum cardiomyopathy)
    – heart failure of any aetiology
  ◼ Valvular heart disease:
    – including previous valve replacement
  ◼ Congenital cardiac lesions:
    – including following surgical correction of congenital lesion
  ◼ Arrhythmia:
    – cardiac conduction defects and arrhythmias
    – history of previous cardiac ablation
    – inherited arrhythmia syndromes
  ◼ Coronary artery diseases, including:
    – atherosclerotic disease
    – spontaneous coronary artery dissection
    – congenital coronary anomalies or arteriovenous malformations
  ◼ Chronic systemic hypertension
  ◼ Pulmonary hypertension
  ◼ Aortic disease
  ◼ Connective tissue disease:
    – heritable collagen, elastin, or glycosaminoglycan disorders
    – autoimmune connective tissue diseases and vasculitides
  ◼ Thromboembolic disease
  ◼ Carriers of (pathogenic) genetic variants associated with cardiovascular conditions
  ◼ Other relevant long-term cardiovascular diagnoses as determined by the study investigators

For those enrolling as participants without cardiovascular disease (controls):

○ Currently pregnant (with pregnancy defined as a positive pregnancy test and/or ultrasound confirmed pregnancy)
○ No diagnosed cardiovascular disease, as defined by the conditions listed above

**Exclusion criteria**

○ Patients who lack capacity to consent
○ People who do not have access to the Internet and/or who are unable to provide an email address for study correspondence
○ Although vulnerable groups (e.g. prisoners, those in dependent relationships) are not ineligible to participate, clinician discretion will be advised when considering whether to signpost potential participants to the registry where such vulnerabilities may be present

### Recruitment and consent

Eligible participants will be directed to the PREG-HEART online platform via multiple routes, including social media campaigns, patient charities and support groups, national and local media promotion, referral from clinicians in relevant specialties (obstetrics, cardiology, anaesthetics, clinical genetics, general practice, and others), and display of study information to female participants in the Heart Hive registry. Clinician awareness will be raised through dissemination across local and national professional networks.

On accessing the online platform (https://thehearthive.org/preg-heart), potential participants will complete screening questions to confirm eligibility. Those eligible will be provided with detailed participant information online and will complete a step-by-step electronic consent process. This includes multi-page walkthroughs and comprehension checks to ensure understanding before enrolment.

Participant information materials (provided in Supplementary File 1) will explain the use of all data and acknowledge that participants can opt out of completing any questions or providing consent for data linkage or storage and use of leftover blood from clinical samples. It also explains how to withdraw from the study should they wish.

### Study procedures

#### Baseline evaluations

Upon registration, participants will be requested to enter socio-demographic details and health data relevant to their cardiovascular diagnosis and current/previous pregnancies via an online questionnaire. Data collected includes socio-demographic and lifestyle information, medical, surgical, and obstetric history, family history of cardiac disease, medications, ECG, echocardiography, cardiac MRI, and other investigations participants may undergo as part of their clinical care. The baseline questionnaire we will use for data collection is included in Appendix 2.

To improve completeness and accuracy of collected clinical data and reduce burden on participants associated with data entry, we will request that participants send (to a secure NHS study email address) copies of clinic letters and results of imaging, genetic, biochemical, or histopathological investigations which pertain to their cardiovascular diagnosis or pregnancy, where these are available. These documents will be securely uploaded to the PREG-HEART platform by the study team.

#### Follow up and long-term data linkage

In addition to the online questionnaire completed when participants first register on the PREG-HEART platform, we will offer participants a further questionnaire within 6 months of the expected pregnancy due date. This will collect information about obstetric and neonatal outcomes. The follow up questionnaire we will use for data collection is included in Appendix 3.

Consent will also be sought for collection of follow-up data over their lifetime from NHS digital and national registries (e.g. Office for National Statistics (ONS), Hospital Episodes Statistics (HES), Patient Episode Dataset for Wales (PEDW), Scottish Morbidity Record (SMR02), Diagnostic Imaging Dataset (DID), Maternity Services Data Set (MSDS), Maternity Indicators Dataset (MIDS), Scottish Birth Record (SBR), National Neonatal Research Database (NNRD), Intensive Care National Audit and Research Centre (ICNARC), and equivalent national registries, as well as further linkage to datasets through the British Heart Foundation Data Science Centre www.bhfdatasciencecentre.org/for-researchers/datasets and Clinical Practice Research Datalink (CPRD) https://www.cprd.com) and from medical records (e.g. primary care data via Vision www.visionhealth.co.uk, Emis www.emishealth.com, SystmOne www.tpp-uk.com/products/systmone, maternity and neonatal data via BadgerNet Maternity www.systemc.com/our-solutions/healthcare/maternity-neonatal, K2 www.k2ms.com, TrakCare for Maternity www.intersystems.com/uk/products/trakcare, and equivalent electronic health and maternity care records). This data linkage aspect of the PREG-HEART study is dependent on future funding.

#### Biosamples

Consent will be sought from participants to collect and store leftover blood from routinely collected clinical samples in pregnancy (which would otherwise be discarded), for unspecified future research use. We will also seek consent to recontact participants in future to invite them to provide further biosamples (blood, saliva, and/or urine).

In the initial phase of this study, existing local biobank infrastructure will be utilised for processing and storage of blood leftover from routinely collected clinical samples. In future phases of the study and as additional funding is secured, biosamples will be transported to a central secure facility for processing and storage.

Collected biosamples may undergo genomic, transcriptomic, proteomic, lipidomic, metabolomic and biochemical analyses, dependent on future funding. Participants provide consent for sequencing up to the level of the whole genome. Data generated from the samples will be linked with demographic and clinical data.

### Substudies

Participants will be asked to provide their consent to be contacted regarding participating in additional ethically approved research studies, focusing on specific aspects of cardiovascular disease in pregnancy. These studies may be led by Imperial College London or collaborating academic and healthcare centres. Specific consent would be obtained from patients for these additional studies, following a clear explanation of any risks involved.

### Data flow and management

Consent forms and data collected in study questionnaires will be entered via the Heart Hive, a secure online web portal built in collaboration with the Broad Clinical Labs (a subsidiary of the Broad Institute, a not-for-profit research institute) on the Juniper data platform. This data will be stored, along with copies of clinic letters and investigation reports submitted by participants, on secure servers in accordance with a data processing agreement with Imperial College London (the data controller).

Further funding will be sought to enable data linkage with NHS digital and national registries, allowing incorporation of additional patient-level information from maternity services, primary care, community dispensing, hospital episode and death statistics. Linked data will be stored in an accredited Trusted Research Environment in collaboration with the BHF Data Science Centre. The BHF Data Science Centre is part of Health Data Research UK, the UK’s Institute for Health data science, and will support coordinated data storage, access, sharing, analysis, and linkage in line with best practices in data management, security and governance.

### Data analysis

#### Pilot study

To evaluate the feasibility and representativeness of participant self-enrolment to our pilot, we will assess recruitment to the PREG-HEART platform at 6 months. We will report numbers of participants enrolled, completion rates of baseline questionnaires, proportion consenting to provide biosamples, the self-reported primary cardiovascular diagnoses of enrolled participants, and geographical, socio-economic, and ethnic distributions.

We will compare participants’ self-reported cardiovascular to estimated UK prevalences. Our group estimates the UK prevalence of heart disease pre-dating pregnancy to be at least 1% (20,475 out of 1,967,250 pregnancies in England between 1^st^ November 2019 and 30^th^ June 2024)(7). Of these, 1,520 women had heart failure and 730 had cardiomyopathy, identified using electronic health record coding data. Based on these figures, we would consider enrolling 25 participants with heart failure or cardiomyopathy (10% of cases over 6 months) during the pilot phase as evidence of feasibility.

We will report the geographical, socio-economic, and ethnic diversity of our enrolled cohort with comparison to that of the UK general pregnant population. This will allow us to identify underrepresented groups and thus will inform our approaches to recruitment for the larger scaled-up study.

#### Future work

Alongside future scale-up of the PREG-HEART platform, we will report outcome data for the cohort collected through follow-up questionnaires and data linkage, on an approximately annual basis. Reported outcomes will include the incidence of major adverse cardiovascular events (MACE), including cardiovascular death, arrhythmic and heart failure events, stroke, atrial fibrillation. Adverse obstetric and neonatal outcomes reported will include maternal death, stillbirth or neonatal death, congenital anomaly, preterm delivery, fetal growth restriction, neonatal unit admission, antenatal or postpartum haemorrhage, and maternal hypertensive disease. Outcomes will be stratified according to participant socio-demographic status and ethnicity, and regional and institutional variations in outcomes will be reported.

### Patient and public involvement and engagement

Women with lived experience of cardiovascular disease in pregnancy have been involved as project partners in all aspects of the study from inception including setting research questions, study design, protocol development, design of patient surveys and all other patient facing materials, development and testing of the online platform, co-authorship of this paper, and planning the dissemination strategy for the registry.

A dedicated patient advisory group, including women with lived experience of cardiovascular disease in pregnancy, will provide ongoing input into strategies for recruitment, further development of the registry, data analysis, and dissemination of findings.

All patient partners are compensated for their time in line with NIHR feasibility guidance.

## Ethics and Dissemination

This study involves human participants and was approved by the West Midlands Solihull Ethics Committee (IRAS project ID: 349694) (REC reference: 25/WM/0142).

Results from the PREG-HEART study will be presented at conferences, published in peer-reviewed academic journals, and be made available on the study’s website (https://thehearthive.org/preg-heart).

## Conclusion

In this pilot study, we aim to demonstrate the feasibility of the PREG-HEART platform for direct-to-patient recruitment of women with cardiovascular disease in pregnancy, together with healthy controls. Future work will further develop this platform through large-scale recruitment, long-term linkage to health outcomes, and integration of a bioresource for multiomic research. This will create an enduring resource for epidemiological analyses, mechanistic discovery, and interventional studies to address critical gaps in evidence on maternal cardiovascular health.

Our approach has the potential to overcome long-standing barriers to participation, generate representative real-world data, and create a resource that is accessible across disciplines and institutions. In doing so, PREG-HEART aims to build national research capacity, advance risk prediction and biomarker development, and support clinical trials that can inform practice and policy. Ultimately, the study seeks to improve outcomes and reduce inequalities for women affected by cardiovascular disease.

## Data Availability

All data produced in the present study are available upon reasonable request to the authors

## Authors’ Contributions

All authors inputted to study design and are involved with study delivery. The manuscript was drafted by KH and RB, and all authors provided critical input and review.

