## Appendices for "Rationale and design of the PREGnancy, HEART Health and Cardiovascular Disease (PREG-HEART) Cohort Study"

**Appendix 1: Membership of NIHR-BHF Cardiovascular Partnership Maternal Cardiovascular Health Theme working group**

Prof. Kate Bramham, King's College London

Rachel Buchan, Imperial College London, Royal Brompton and Harefield Hospitals, Guy’s and St Thomas’ NHS Trust

Rachel Cartwright, Imperial College London

Dr Matthew Cauldwell, St George's University Hospitals NHS Foundation Trust

Prof. Tim Chico, Sheffield Teaching Hospitals NHS Foundation Trust, University of Sheffield and BHF Data Science Centre

Dr Paul Clift, Queen Elizabeth Hospital Birmingham

Dr Simon Fisher, NIHR-BHF Cardiovascular Partnership and University of Oxford

Lisa Godden, Patient Representative

Dr Kathryn Hunt, University of Edinburgh

Rebecca Jarman, Research Midwife and NIHR Clinical Research Network South London

Prof. Mark Johnson, Chelsea and Westminster Hospital NHS Foundation Trust

Dr Jamie Kitt, University of Oxford and Oxford University Hospitals NHS Foundation Trust

Prof. Deborah Lawlor, University of Bristol

Prof. Paul Leeson, University of Oxford and Oxford University Hospitals NHS Foundation Trust

Dr Antonio de Marvao, King's College London and Imperial College London

Prof. Edward Mullins, Imperial College London

Prof. Jenny Myers, University of Manchester and St Mary’s Hospital

Dr Laura Ormesher, University of Manchester and Manchester University NHS Foundation Trust

Dr Elena Raffetti, University of Cambridge and Uppsala University

Prof. Rebecca Reynolds, University of Edinburgh

Dr Nora Rossberg, Imperial College London

Claire Sheppard, Patient Representative

Prof. Bee Tan, University of Leicester and University Hospitals Leicester NHS Trust

Dr Upasana Tayal, Imperial College London, Royal Brompton and Harefield Hospitals, Guy’s and St Thomas’ NHS Trust

Prof. Basky Thilaganathan, St. George's University Hospitals NHS Foundation Trust

Prof. David Williams, University College London Hospital

Prof. Catherine Williamson, Imperial College London

**Appendix 2: Baseline questionnaire**

Identifying information:

1. Are you registered in the UK with a GP?
2. If yes, where are you registered (England/Wales/Scotland/Northern Ireland/Other)
3. Please enter your NHS number (England or Wales)/CHI number (Scotland)/H&C number (Northern Ireland)

Questions about heart or blood vessel condition:

1. Which of the following types of heart or blood vessel conditions have you been diagnosed with? *Please select (you can choose more than one option if needed)*
   - cardiomyopathy or heart failure – this is disease affecting the heart muscle which reduces the pumping function of the chambers called ventricles
   - heart valve problems (narrowed or leaky valves, sometimes called stenosis or regurgitation)
   - congenital heart disease – this means a problem with the structure of the heart that you were born with, including if you needed surgery on your heart as a child
   - cardiac arrythmia – this includes any disease that causes an abnormal heart rhythm
   - coronary artery disease – this is a problem affecting the blood supply to the heart, caused by blocked or narrowed vessels supplying the heart
   - hypertension – high blood pressure
   - pulmonary hypertension – increased pressure specifically in the blood vessels carrying blood towards your lungs
   - aortic disease – any problem affecting the main blood vessel carrying blood away from the heart (the aorta)
   - connective tissue disease – these disorders can be inherited (genetic, e.g. Marfan Syndrome, Ehlers-Danlos syndrome) or have environmental or unknown causes (e.g. SLE/lupus, vasculitis, amyloidosis)
   - thromboembolism – a blood clot in a major blood vessel, e.g. affecting the lungs, legs, brain or eyes
   - other – *please specify*

Please give the specific name of your condition (ideally from any letters from your doctors), if you know this:

2. When was your heart or blood vessel condition first diagnosed?
   - Before my most recent/current pregnancy
   - During my most recent/current pregnancy, or in the six months after giving birth
3. Who manages your heart or blood vessel condition?
   - Please enter the name of your cardiologist if you know this
   - Please enter the name of the hospital
4. Have you ever had any surgery on your heart? *Please enter the date of this surgery, and any details about the type of surgery if you are able.*
5. An echocardiogram, or “echo” is a type of ultrasound scan which looks at the heart. It is carried out by placing a small probe on your chest to create a moving image of your heart as it is beating **INSERT PICTURE**

Have you ever had an echocardiogram?

Please enter the approximate date of your last echocardiogram

Please enter the name of the hospital where you had this scan

Do you have a copy of your echocardiogram report? Please email this to the study team if you have it available.

6. A cardiac MRI scan is a non-invasive test that uses an MRI machine to create magnetic and radio waves to create clear pictures showing the inside of your heart. Unlike an X-ray, an MRI scan does not use radiation. You lie on a bed, which moves inside a cylindrical scanner

Have you ever had a cardiac MRI (CMR) scan?

Please enter the approximate date of your last cardiac MRI scan

Please enter the name of the hospital where you had this scan

Do you have a copy of your cardiac MRI scan report? Please email this to the study team if you have it available.

7. Please email to the study team any recent clinic letters from appointments with your cardiology team during the last three years, if you have these available.
8. For some heart conditions, genetic testing to identify possible causes is becoming more common.

Have you had genetic testing for a heart or blood vessel condition?

Which hospital did you have your genetic test at?

What was the rest of your genetic test?

- Definite genetic cause identified
- A genetic variant of uncertain significance – might be the cause of my condition, but further testing needed
- Normal, no genetic cause found
- Still awaiting result
- Don’t know

Please enter the name of the gene identified as the cause of your heart or blood vessel condition.

Do you have a copy of your genetic test result? Please email this to the study team if you have it available.

9. Do/did any of your parents or siblings have a heart condition? *Please specify which relative(s), and what condition(s) they have/had.*

Questions about current/most recent pregnancy

1. Which hospital(s) provides/provided your pregnancy (antenatal and labour) care during your current/most recent pregnancy?

Research participation:

1. Have you ever taken part in other medical or health research? *If yes, please give the name of each study you have participated in.*

Sociodemographic questions:

1. Which of the following best describes your ethnicity?
   - White - British
   - White - Irish
   - White - Any other White background
   - Mixed - White and Black Caribbean
   - Mixed - White and Black African
   - Mixed - White and Asian
   - Mixed - Any other mixed background
   - Asian or Asian British - Indian
   - Asian or Asian British - Pakistani
   - Asian or Asian British - Bangladeshi
   - Asian or Asian British - Any other Asian background
   - Black or Black British - Caribbean
   - Black or Black British - African
   - Black or Black British - Any other Black background
   - Other Ethnic Groups - Chinese
   - Other Ethnic Groups - Any other ethnic group
   - I prefer not to answer
2. Is the gender you identify with the same as your sex registered at birth?
   - Yes
   - No
   - I prefer not to answer
3. What is your postcode?

**Appendix 3: Follow up questionnaire**

1. What was the outcome of your recent pregnancy
   - miscarriage – this is the loss of a pregnancy before 23 completed weeks of pregnancy
   - termination of pregnancy
   - ectopic pregnancy – this is when a fertilised egg implants outside of the womb, usually in one of the fallopian tubes
   - stillbirth – this is where a baby dies in the womb after 24 weeks of pregnancy
   - birth of a live baby
2. During your pregnancy or following the birth, was your baby diagnosed with any structural abnormality (sometimes called a “congenital anomaly” or “birth defect”) or genetic condition? *Please specify*
3. During your pregnancy, or in the first six months following the end the pregnancy, were you diagnosed with any of the following:
   - gestational hypertension or pre-eclampsia – these are problems where your blood pressure becomes too high during or soon after the end of pregnancy
   - obstetric cholestasis – this is a condition affecting the liver which can occur in pregnancy
   - gestational diabetes – this is high blood sugar that develops during pregnancy and usually resolves after giving birth
   - postnatal depression
   - Any other condition relating to pregnancy – *please specify*
4. At what gestation (number of completed weeks of pregnancy) was your baby born?
5. Was your labour started off, e.g. with medications, a device that is put into the neck of your womb, or by a doctor or midwife breaking your waters before contractions started (this is called “induction of labour”)?
6. How was your baby born?
   - Vaginally without needing any instruments
   - Vaginally, with forceps or a vacuum cup (“sometimes called a “kiwi” or “ventouse”)
   - Caesarean section
7. During or after the birth, did you lose more than one litre of blood or need a blood transfusion?
8. What was the approximate weight of your baby when they were born?
9. Did your baby need to be admitted to a neonatal intensive care unit after they were born?
10. Is your baby still alive? *If no – what age were they when they died?*
11. Was your baby diagnosed with any long-term health problems during the first six months of their life? *Please specify*
12. Please email to the study team with clinic letters you have from appointments with your cardiology team or obstetrician relating to this pregnancy, if you have these available.
