## supplementary file 1 for "Rationale and design of the PREGnancy, HEART Health and Cardiovascular Disease (PREG-HEART) Cohort Study"

**Participant Information Sheet**

You are being invited to take part in a research study. Before you decide whether to take part, it is important for you to understand why the research is being done and what it will involve. Please take time to read the following information carefully and discuss it with others if you wish.

Ask us if there is anything that is not clear, or if you would like more information – you can contact us at ****. Take time to decide whether you wish to take part.

Thank you for taking the time to read this*.*

### Introduction

### Heart disease is the leading cause of serious illness and death for pregnant women in the UK and can have an important affect on the health of both mother and baby during pregnancy. Despite advancements in medicine, there have been few improvements in this area for many years.

### We want to understand more about how heart disease affects women during and after pregnancy. By collecting information from many women across the UK, we hope to improve care and find better ways to support women with heart disease in pregnancy and their babies.

### What is the purpose of the study?

We are setting up a new research study to learn more about heart health during and after pregnancy. Our first goal is to see if we can build a large, diverse group of women; some with heart conditions and some without, who are pregnant or have recently been pregnant on our secure, online platform.

We want to find out if it’s possible to:

- Recruit and keep participants involved over time
- Collect high-quality health information and collect samples
- Link this data to national health records

We then aim to use this data to:

- understand how heart disease affects pregnancy and how pregnancy affects heart disease
- learn if and how heart conditions are managed across the U.K., and whether there are any differences
- provide an online platform for future research, including studies that may involve collecting biological samples (like blood) to understand more about these conditions.

This is an observational study, which means we will collect information about you and your health over time, but will not change your medical care or treatment.

This study isn’t testing a new treatment or medical procedure. It’s focused on understanding health using information that’s mostly already been collected.

We will collect detailed medical and lifestyle information at the start of the study, ask you to complete health survey data during the course of the study, and may collect blood or saliva samples during the course of the study (subject to funding). We will also gather data collected during routine healthcare visits over time.

### Why have I been invited?

You have been invited to participate because:

- You have been diagnosed with heart disease before, during, or within six months after pregnancy.

OR

- You are currently pregnant without any heart disease and wish to participate as a healthy control.

Heart conditions include (but are not limited to):

- Heart muscle diseases (cardiomyopathies, heart failure)
- Heart valve diseases
- Congenital heart defects (conditions present from birth)
- Heart rhythm problems (arrhythmias)
- Coronary artery disease (problems with blood flow to the heart)
- High blood pressure (hypertension)
- Pulmonary hypertension (high blood pressure in the lungs)
- Aortic diseases (problems with the main artery)
- Connective tissue diseases
- Blood clots (thromboembolic disease)
- Carriers of genetic mutations associated with heart conditions

We will conduct the study in phases. In the first phase we will aim to recruit 200 people. This will be expanded in the next phases.

### Do I have to take part?

Taking part in this research study is entirely optional. It is up to you to decide whether or not to take part. If you do decide to take part, you will be given this information sheet to keep and be asked to complete a consent form.

If you decide to take part, you are free to withdraw at any time and without giving a reason.

A decision to withdraw at any time, or a decision not to take part, will not affect the standard of care you receive from your doctors*.*

If you withdraw from the study, you can choose whether:

- You would like to stop receiving any further study communication (like emails or follow-up questionnaires),
- You would like the study team to stop collecting any further information about you (for example, from medical records or national health registries),
- Or whether you would prefer for all data and samples you’ve already provided to be removed from the study where possible, and for no future data to be collected.

We cannot remove data from research that is underway or has already been done; or remove all records related to you from our databases.

An audit record is needed for our research to be reliable. We will need to confirm that you were once part of the PREG-Heart and then withdrew; This information includes your first name, surname, date of birth, address and contact details.

You can choose to stop receiving any future messages or study updates at any time by updating your profile preferences on the study platform (thehearthive.org), or by contacting the study team directly.

### What will happen to me if I take part?

If you decide to join this research study we won’t change your medical care, but we will ask you to:

- Complete health surveys every year for as long as you are happy to
- Allow us access to your medical records to gather information
- Allow us to access left-over stored samples that may have been collected during your routine NHS care

In the future, we may ask you to provide a blood sample or a saliva sample.

When we collect your data for research, we remove your name and other personal details and replace them with a code. This means researchers can’t identify you directly. Your information is still protected, and only approved people can access it in secure systems. If needed, your data can be linked back to you by a small number of authorised people. We call this data “pseudonymised”.

### What do I have to do?

We won’t be asking you to complete many activities for the study – we will mainly look to collect information about your health from your medical records.

**Heart investigations**

We would like to use data from the heart tests that you have during your usual clinical care. We will store pseudonymised copies of heart tests that you have had in the past, or will have in the future, in our secure research database.  We are particularly interested in digital images of the heart from cardiac MRI or ultrasound (echocardiogram), and digital heart rhythm traces (ECG / electrocardiogram).

We will use tests carried out as part of your routine clinical care. You will not have to have any extra tests.

We may also upload these pseudonymised test results to other study sites based outside the UK, using a unique study ID to label your scan and removing your personal details (name, date of birth).

The cardiac imaging repository will allow researchers to look more deeply into the structure and function of the heart by using the digital data available for research.

**Uploading documents**

We will ask you to upload documents like clinic letters or test results related to your heart condition or pregnancy. This is **optional** - you can still take part even if you do not upload anything.

You can upload photos taken on your phone, scanned copies, or digital versions. Documents can be uploaded during registration or at any later point when you are ready.

**Follow Up**

We would like you to provide updates about your health during the study. We will invite you to complete short annual questionnaires about your health either using the Heart Hive (thehearthive.org) platform or offline (via the post or on the telephone/video call with one of the study team) for as long as you are willing to.

We would like your permission to access your Heart Hive health survey data (if completed) and registration details (e.g. date of birth), and to follow up your medical health from your health records over your lifetime. This would not require any direct contact with you. We may collect information relating to your heart condition from your GP and from any hospitals that you have been treated at.

Information about your health is collected routinely by the NHS for national registries such as the Office for National Statistics and Hospital Episodes Statistics. We will link your study data to the data from national registries e.g. Nicor, SSNAP, Public Health Scotland. These national datasets are managed by national organisations, such as NHS England (formerly NHS Digital). We will collect information about your health, and whether you have experienced any heart problems, complications, or any procedures or investigations related to your heart. We will collect information about your medications and treatments related to your heart from your hospital records, and where available GP records and digital records.

We will also collect information about your baby’s health around the time of birth, such as birthweight, gestation at delivery, and whether the baby needed specialist care. No tests or procedures will be done on your baby, and we will not collect any samples from them. We will also not collect any additional clinical information about your baby beyond the immediate period after birth.

The research team may also ask you if you would like to take part in other related research studies. Your participation in any other research studies would be optional. You would be given detailed information about these to help you decide whether you want to take part in these studies.

**Providing samples**

In the future, we may ask you to provide a sample of blood (or saliva if a blood sample is not feasible).

These samples will be pseudonymised, labelled with a study code and stored securely at the study site.

Blood samples

We may contact you to collect some blood samples from you to extract DNA for genetic analysis and to measure changes in the make-up of the blood that can be useful to track heart health. We will not collect more than 50 ml (3.5 tablespoons) of blood from you, and possibly less depending on your age and weight.

*OR*

Saliva

Where it is not feasible or possible to collect a blood sample, a saliva sample may be collected instead. It is simple to provide a saliva sample, and instructions will be available on the saliva kit that will be provided. This will provide us with similar genetic information that we would have obtained from a blood sample.

**Using Existing Samples**

**Blood samples**

If you have provided blood samples as part of your medical care in pregnancy or in the past, we may wish to access leftover samples to investigate your heart health and other genetic information.

**Other tissue samples**

If you have provided a biopsy or other tissue sample as part of your medical care in the past, or if you provide one in the future, we may wish to access leftover tissue to investigate your heart health and other genetic information.

No payments or travel expenses will be provided for taking part in the study.

### Who will have access to my samples and information?

Your privacy and the confidentiality of your information are extremely important to us. Your samples and data will be stored securely. Data will be stored on an accredited Trusted Research Environment (TRE) which has been approved by Imperial College London.

Only authorized members of the research team will have access to your identifiable information, and only when it is necessary for the study. They are bound by strict confidentiality agreements. We will send participant details to a trusted third party (e.g. Digital Health and Care Wales) and to NHS England to perform linkage to national datasets.

We will assign a unique study identification number to your samples and data. This means your personal details, such as your name and NHS number, will be removed and replaced with this code. Researchers analysing the data will not be able to identify you personally.

With your consent, we may share your pseudonymised data and samples with approved researchers within the UK and internationally. This sharing is done to enhance scientific understanding and improve care for women with heart disease in pregnancy.

Your pseudonymised samples and data may be used in future ethically approved research studies related to heart disease and pregnancy. Any such research will require additional ethical approval, and your identity will remain protected.

### What are the possible benefits of taking part?

We do not expect that you will directly benefit from taking part in this research study, though some research participants report psychological benefits from helping to advance medical understanding of their condition.

We anticipate that others with heart disease in pregnancy may benefit in the future from what we learn in this study.

### What are the possible disadvantages and risks of taking part?

The study primarily uses data collected during routine healthcare, with relatively few additional risks. Data protection measures to protect your privacy are discussed below. We will assign you a study ID number and will not use your name, address or date of birth to label your samples or data.

Any genetic data that is generated will also be labelled with your study ID. As your genetic data is unique to you there is a very small risk that it could be used to identify you, but there are many safeguards in place to protect your data, and there would be no benefit or motivation for anyone to ever attempt do this. Researchers will not attempt to identify you.

We may collect additional blood samples. The physical risks of this are minimal. It is possible that you might experience some discomfort, bleeding or bruising following the collection of blood samples for the study.  These effects are generally mild and short lived.

Analyses of genetic data are not expected to generate results that will be useful to the care of individual participants, and so genetic results will not be returned to participants.  There is therefore no risk of harm from unexpected genetic secondary findings. We do not expect any incidental findings to arise from the current research, as we will be using medical data that was collected during your usual care.

### What if new information becomes available?

Sometimes during the course of a research project, new information becomes available about the conditions that are studied. If this happens, the study team may tell you about it. As this study does not change your medical care in any way, your research participation is unlikely to be affected by this.

Also, on receiving relevant new information, the research team might consider it to be in your best interests to withdraw you from the study. The research team will contact you to discuss this.

If we discover any new findings relevant to your condition, we may contact your clinical team or ask you to provide the details of your clinical team. We are unlikely to find new information that would not be found during your routine medical care.

### What if something goes wrong?

Imperial College London holds insurance policies which apply to this study. If you experience harm or injury as a result of taking part in this study, you will be eligible to claim compensation without having to prove that Imperial College is at fault. This does not affect your legal rights to seek compensation.

If you are harmed due to someone's negligence, then you may have grounds for a legal action. Regardless of this, if you wish to complain, or have any concerns about any aspect of the way you have been treated during the course of this study then you should immediately inform the Principal Investigators (Dr Upasana Tayal and Dr Antonio de Marvao, email ****). If you are still not satisfied with the response, you may contact the Imperial College Research Governance and Integrity Team on.

### What will happen to my samples?

Your blood/saliva, optional tissue and DNA samples will be stored in a secure location in the Imperial study site. Stored samples will only be accessible to approved researchers.  Your samples may be used for genetic analyses.  Your blood/saliva samples and DNA may be processed outside of the United Kingdom, depending on where the best or most cost-effective technology is available, but no additional personally identifiable information will be shared, so the samples cannot be traced back to you outside the research team.

Your samples and results from tests on your samples may be shared with other researchers, including those based outside of the United Kingdom. Any research that uses your samples will need to be ethically approved.

At the end of the project if any of your samples remain, they will be stored by the research team.  They may be used in other research projects. An ethics committee will review and approve any new projects that your samples may be used for.  If your samples are not used for any other research projects, they may be disposed of according to applicable guidelines.

### Will my taking part in this study be kept confidential?

All information which is collected about you during the course of the research will be kept strictly confidential.  Everyone handling your personal and medical details will be bound by a professional duty to protect your privacy. When research studies are published from this project, they will not contain your identifiable details (name, date of birth, etc) and it will not be possible for anyone else to identify who you are.

Study staff will assign a study ID number to your samples and cardiac imaging files. Your samples and, if available, your cardiac imaging will not be labelled with any information that could identify you. The staff will use the study ID to connect your sample to your health information that is stored in a computer database. The computer database is protected with a password. Only study staff will know the password.

Only the UK research team members with appropriate permissions will have access to the key linking your study ID to your personal information. This will only be used when we need to update your information or contact you. Researchers will not attempt to identify you when analysing study data.

Your genetic and genomic results may be stored on secure computer systems at Imperial College and/or other collaborating research centres.  These results will be labelled with your study ID.  This information will not include your identifiable information (name, date of birth, etc.).

With your permission we may ask your GP to provide relevant information about your cardiovascular health to us.

### Will I receive any genetic results?

We will not provide feedback on the results of research on your blood or saliva samples.

If your clinical team requests access to your genetic results to inform your clinical care, we are happy to provide relevant data to them with the understanding that any results will need to be confirmed by a diagnostic laboratory, however, we will ask for your consent to do so. Requests in the first instance from your clinician may be made to.

We will only be looking at genetic variants related to cardiovascular disease and will not be screening for other genetics results in different disease types. We will not report back any genetic findings relating to other conditions.

### What will happen if I lose the ability to make decisions about my participation in the study?

### If, during the course of the study, you lose the ability to make decisions about your participation (lose capacity), the following will apply:

### Your care and well-being will always remain the priority. If you lose capacity, we will review your continued involvement in the study. We will consult with a suitable person acting in your best interests, such as a relative, close friend, or an appointed legal representative (known as a consultee or personal/nominated representative).

### If your representative believes that continuing in the study is not in your best interests, or if you show any signs of distress or objection, you will be withdrawn from the study, and this will not affect your medical care in any way.

### If it is appropriate for you to remain in the study, your representative will act on your behalf for study-related decisions as required by law. You may be withdrawn from the study at any time if your representative requests it.

### Any samples or data collected up to the point you lose capacity may still be used in line with the study protocol, unless your representative asks for them to be withdrawn.

### What will happen to the results of the research study?

To increase the benefit from your participation in this study, we may share our research findings with other researchers.  Results from research on your samples and health information may be published on controlled access databases, including genetic research.  Controlled access databases mean that only researchers who apply for and get permission to use the information for a specific research project can access the information.  Your genetic and health information would not be labelled with your name, date of birth, or other information that could be used to identify them.  Researchers approved to access information in the database will agree not to attempt to identify you.

Sharing research results on controlled access databases will allow researchers from other organisations to use your information to study genetic causes of diseases.  These databases may be located in countries outside of the United Kingdom and European Union.  Data protection laws in other countries may not offer the same level of privacy protection as those in the UK, but none of your identifiable information (name, date of birth, etc) will be included on these databases.

At the end of the project all the research results will be gathered together and analysed.  The researchers have a professional responsibility to publish their findings, however your identity will not be revealed. Most research is published in the medical press – if you are interested in knowing the overall results of the study, ask the research team about this.

### Who is organising and funding the research?

The research project is being organised by the research team at Imperial College London and working with partner hospitals in collaboration with an international study team.

The research is being funded by the National Institute for Health and Care Research and the British Heart Foundation.

### Who has reviewed the study?

This study was given a favourable ethical opinion for conduct in the NHS by West Midlands Solihull Research Ethics Service.

### Contact for Further Information

Please contact the study team on the following contact details:

### GDPR and Imperial College Privacy Notices

**How will we use information about you?**

Imperial College London is the sponsor for this study and will act as the data controller for for this study. Being a Data Controller means that we are responsible for looking after your information and using it appropriately plus are responsible for explaining this to you. Imperial College London will keep your personal data for:

- 10 years after the study has finished in relation to data subject consent forms.
- 10 years after the study has completed in relation to primary research data.

 The study is expected to finish in August 2029. We may extend the study further if we secure further funding and ethical approval.

For more information / confirmation regarding the end date please contact the study team, see ‘WHERE CAN YOU FIND OUT MORE ABOUT HOW YOUR INFORMATION IS USED’ for contact information

We will need to use information from you, your medical records and your GP for this research project.

This information will include your:

NHS number

Name

Date of Birth

Contact details

People within the College and study team will use this information to do the research, to check your records and make sure that the research is being done properly, and to check that information held (such as contact details) is accurate.

People who do not need to know who you are will not be able to see your name or contact details. Your data will have a code number instead.

We will keep all information about you safe and secure.

- Data management plans have been created and reviewed in line with Imperial’s Information Governance Policy Framework. This covers the collection, movement, processing and storage of the data.
- Data to be stored in a dedicated secure environment which underpins security measures.
- Data will be stored in ISO 27001 certified and/or Cyber Essentials accredited environment
- Robust pseudonymisation has been implemented to prevent identification
- Access controls have been implemented to ensure only key personnel can access the data

Some of your information will be sent to the USA. They must follow our rules about keeping your information safe.

Once we have finished the study, we will keep some of the data so we can check the results. We will write our reports in a way that no-one can work out that you took part in the study.

As a university we use personally-identifiable information to conduct research to improve health care and services. As a publicly-funded organisation, we have to ensure that it is in the public interest when we use personally-identifiable information from people who have agreed to take part in research. This means that when you agree to take part in a research study, we will use your data in the ways needed to conduct and analyse the research study. Our legal basis for using your information under the General Data Protection Regulation (GDPR) and the Data Protection Act 2018, is as follows:

• Imperial College London - “performance of a task carried out in the public interest”; Health and care research should serve the public interest, which means that we have to demonstrate that our research serves the interests of society as a whole. We do this by following the UK Policy Framework for Health and Social Care Research

Where special category personal information is involved (most commonly health data, biometric data i.e. finger prints or facial recognition and genetic data, racial and ethnic data etc.), Imperial College London relies on “scientific or historical research purposes” or “statistical purposes”.

**International transfers**

We may share data about you outside the UK for research related purposes to:

- Where necessary to provide access to a data processor/service provider who will utilise your personal data as instructed by us.
- Where necessary to share with a third party organisation / collaborator (as listed) who are also involved in the study.
- Where data has been collected from outside the UK and requires additional transfer(s) as part of the study activity.

If this happens, we will only share the data that is needed. We will also make sure you can’t be identified from the data that is shared where possible. This may not be possible under certain circumstances – for instance, if you have a rare illness, it may still be possible to identify you. If your data is shared outside the UK, it will be with the following sorts of organisations:

- The Broad Institute, USA – hosting the Heart Hive website

We will make sure your data is protected. Anyone who accesses your data outside the UK must do what we tell them so that your data has a similar level of protection as it does under UK law. We will make sure your data is safe outside the UK by doing the following

- we use specific contracts which stipulates that personal data must maintain the same level of protection when outside the UK as it has within the UK. For further details [visit the Information Commissioner’s Office (ICO) website](https://ico.org.uk/for-organisations/uk-gdpr-guidance-and-resources/international-transfers/) - [www.ico.org.uk](http://www.ico.org.uk)
- we do not allow those who access your data outside the UK to use it for anything other than what our written contract with them says
- we need other organisations to have appropriate security measures to protect your data which are consistent with the data security and confidentiality obligations we have. This includes having appropriate measures to protect your data against accidental loss and unauthorised access, use, changes or sharing
- we have procedures in place to deal with any suspected personal data breach. For further details about UK breach reporting rules [visit the Information Commissioner's Office (ICO) website](https://ico.org.uk/for-organisations/report-a-breach) - [Personal data breaches: a guide | ICO](https://ico.org.uk/for-organisations/report-a-breach/personal-data-breach/personal-data-breaches-a-guide/#whendowe)

**Sharing your information with others**

We will only share your personal data with certain third parties for the purposes referred to in this participant information sheet, and by relying on the legal basis for processing your data as set out above:

- Other Imperial College London employees (including staff involved directly with the research study or as part of certain secondary activities which may include support functions, internal audits, ensuring accuracy of contact details etc.), Imperial College London agents, contractors and service providers (for example, suppliers of printing and mailing services, email communication services or web services, or suppliers who help us carry out any of the activities described above). Our third party service providers are required to enter into data processing agreements with us. We only permit them to process your personal data for specified purposes and in accordance with our policies.
- the following Research Collaborators / Partners in the study

-The BHF Data Science Centre – we aim to work with the BHF Data Science Centre (or a similar organisation) to build a Trusted Research Environment, to link and store NHS England Data collected from participants in a secure way. This data will only be accessible to approved users and cannot leave the secure environment. We share identifiable details to link this data up, but these are removed before being stored in the secure environment, so users only see a unique study ID.

**Potential use of study data for future research**

When you agree to take part in a research study, the information collected either as part of the study or in preparation for the study (such as contact details) may, if you consent, be provided to researchers running other research studies at Imperial College London and in other organisations which may be universities or organisations involved in research in this country or abroad. Your information will only be used to conduct research in accordance with legislation including the GDPR and the [UK Policy Framework for Health and Social Care Research](https://www.hra.nhs.uk/planning-and-improving-research/policies-standards-legislation/uk-policy-framework-health-social-care-research/).

This information will not identify you and will not be combined with other information in a way that could identify you, used against you or used to make decisions about you.

**Commercialisation**

Samples or data from the study may also be provided to organisations not named in this participant information sheet, e.g. commercial organisations or non-commercial organisations for the purposes of undertaking the current study, future research studies or commercial purposes such as development by a company of a new test, product or treatment. We will ensure your name and any identifying details will NOT be given to these third parties, instead you will be identified by a unique study ID with any sample / data analysis having the potential to generate ‘personal data’.

Aggregated (combined) or pseudonymised data sets (all identifying information is removed) may also be created using your data (in a way which does not identify you individually) and be used for such research or commercial purposes where the purposes align to relevant legislation (including the GDPR) and wider aims of the study. Your data will not be shared with a commercial organisation for marketing purposes.

**What are your choices about how your information is used?**

- You can stop being part of the study at any time, without giving a reason, but we will keep information about you that we already have.
- You have the right to ask us to remove, change or delete data we hold about you for the purposes of the study. We might not always be able to do this if it means we cannot use your data to do the research. If so, we will tell you
- if follow up data will be collected after withdrawal: If you choose to stop taking part in the study, we would like to continue collecting information about your health from [central NHS records / your hospital / your GP]. If you do not want this to happen, tell us and we will stop

**Where can you find out more about how your information is used?**

You can find out more about how we use your information.

·  	at [www.hra.nhs.uk/information-about-patients/](https://www.hra.nhs.uk/information-about-patients/).

·  	by asking one of the research team.

**Complaint**

Following our response, if you are not satisfied please contact Imperial College London’s Data Protection Officer via email at, via telephone on 020 7594 3502 and/or via post at Imperial College London, Data Protection Officer, Faculty Building Level 4, London SW7 2AZ

If you remain unsatisfied with our response or believe we are processing your personal data in a way that is not lawful you can complain to the Information Commissioner’s Office (ICO)- via [www.ico.org.uk](http://www.ico.org.uk). Please note the ICO does recommend that you seek to resolve matters with the data controller (us) first before involving them.

**Thank you for considering taking part in this study!**

You can access a copy of this sheet from your Heart Hive dashboard.
